# Functional connectivity of stimulus-evoked brain responses to natural speech in post-stroke aphasia

**DOI:** 10.1101/2024.01.15.24301324

**Authors:** Ramtin Mehraram, Pieter De Clercq, Jill Kries, Maaike Vandermosten, Tom Francart

## Abstract

One out of three stroke-patients develop language processing impairment known as aphasia. The need for ecological validity of the existing diagnostic tools motivates research on biomarkers, such as stimulus-evoked brain responses. With the aim of enhancing the physiological interpretation of the latter, we used EEG to investigate how functional brain network patterns associated with the neural response to natural speech are affected in persons with post-stroke chronic aphasia.

EEG was recorded from 24 healthy controls and 40 persons with aphasia while they listened to a story. Stimulus-evoked brain responses at all scalp regions were measured as neural envelope tracking in the delta (0.5-4 Hz), theta (4-8 Hz) and low-gamma bands (30-49 Hz) using mutual information. Functional connectivity between neural-tracking signals was measured, and the Network-Based Statistics toolbox was used to: 1) assess the added value of the neural tracking vs EEG time series, 2) test between-group differences and 3) investigate any association with language performance in aphasia. Graph theory was also used to investigate topological alterations in aphasia.

Functional connectivity was higher when assessed from neural tracking compared to EEG time series. Persons with aphasia showed weaker low-gamma-band left-hemispheric connectivity, and graph theory-based results showed a greater network segregation and higher region-specific node strength. Aphasia also exhibited a correlation between delta-band connectivity within the left pre-frontal region and language performance.

We demonstrated the added value of neural tracking when investigating functional brain networks associated with natural speech processing in post-stroke aphasia. Its higher sensitivity to language-related brain circuits favours its use as informative biomarker for the assessment of aphasia.

## 1. Introduction

Aphasia is an acquired language-related disorder most often caused by a stroke. On average, one out of three stroke patients develop impairments in language processing,^1^ with significant inter-individual variability in terms of symptoms and pathology.^2^ Behavioral assessment has limited validation^3,4^ and might be capturing comorbid disorders. Hence, it may benefit from supportive biomarkers.

According to task-based and resting-state neuroimaging studies, stroke-related aphasia is associated with alteration in the functional organization of the brain activity, as assessed with both fMRI and EEG.^5^ Depending on the stage and severity of the pathology, post-stroke aphasia exhibits affected functional integration and connectivity distribution of the brain network, with a shift of activation towards lower frequency bands within the lesioned hemisphere and increased cortical activation over the contra-lesional hemisphere, likely associated with compensatory mechanisms.^6–11^ In recent research involving part of the participant cohort of the present study, we used EEG and investigated functional connectivity alterations in persons with chronic post-stroke aphasia (PWA) during natural speech listening.^9^ We found that PWA exhibit higher interhemispheric connectivity compared to healthy controls (HC) within the theta-band (4.5-7 Hz) network and weaker widespread connectivity within the low-gamma band (30.5-49 Hz). These results support the hypothesis of ongoing compensatory processes in PWA associated with recovery and resonate with pathological reorganization of language- and auditory-related functional networks^12,13^ towards a more functionally centered architecture.^9^

Language processing and related disorders can also be investigated through the analysis of neural tracking of natural speech. This is performed by quantitatively assessing the statistical relationship between the EEG-recorded neural signal and the speech stimulus. In fact, the envelope of natural speech has been shown to be informative enough to allow for a good level of comprehension,^14,15^ and its neural tracking is reportedly an effective marker of speech understanding and intelligibility.^16–19^ Interestingly, neural tracking is also associated with a functional connectome, i.e., the neural response to the envelope of natural speech is synchronized across brain regions and can be defined as a functional network.^20^ The analysis of such network may provide further insights into the functional and structural processes associated with the processing of natural speech, although its added value as a biomarker compared to the connectome associated with EEG time series has not been investigated yet.

Despite the existing evidence of neural envelope tracking as a biomarker for diverse language disorders,^21–24^ research on post-stroke aphasia to date remains limited. In our most recent works, we used temporal response functions (TRF)^25^ and temporal mutual information functions (TMIF)^26,27^ as measures of neural tracking. The first consists of a linear model, where weights represent the amplitude of the brain response to the stimulus at different latencies; the latter is a statistical approach also capturing non-linear relationships between the EEG and the stimulus, and provides a more thorough assessment of neural tracking.^26^ Using TMIFs, we reported reduced neural envelope tracking in PWA compared with HC within fronto-central, parietal and posterior scalp regions. Affected frequency bands included delta (0.5 – 4 Hz), theta (4 – 8 Hz) and low-gamma (30 – 49 Hz), suggesting an impairment of lower-level acoustic and higher-level linguistic structures,^17,28,29^ such as encoding of phonetic features (Giraud & Poeppel, 2012; Gross et al., 2013; Hyafil et al., 2015).

In the present study, we investigate whether the functional connectome associated with natural speech processing is altered in PWA. In a combined manner, we measure the neural envelope tracking of natural speech as TMIF, and we assess the strength of synchronization of the neural tracking between scalp regions. To infer the added value of such an approach, we first compare the functional connectome associated with neural envelope tracking to the network derived from the EEG time series. We then compare the TMIF-connectome between HC and PWA, and perform graph theory analysis to assess between-group differences. Our analyses focuses specifically on the delta (0.5-4 Hz), theta (4-8 Hz) and low-gamma (30-49 Hz) frequency bands, since these reportedly reflect diverse aspects of language processing and its disorders.^9,12,13,27,28^

## 2. Materials and methods

### 2.1. Participants

PWA were recruited within the cohort of participants at the stroke unit of the University Hospital Leuven, Belgium (UZ Leuven). Patients’ screening was performed in the acute stage with the help of speech and language pathologists using a Dutch adaptation of the Language Screening Test (LAST),^31–33^ with a cut-off score for inclusion of 14/15. Only PWA with left-hemispheric or bilateral lesion were recruited. For each included patient, the experimental session took place not earlier than six months following the stroke onset (median: 17 months; range: 6-369 months). Recruitment of HC was pursued aiming to match the average age of PWA. All participants had no history of psychiatric or neurodegenerative disorders and gave written informed consent before participation. The study was approved by the Medical Ethical Committee UZ/KU Leuven (S60007) and was performed in accordance with the relevant guidelines and regulations.

Our cohort comprised 24 HC (72 ± 7 years old, 16 males) and 40 PWA (69 ± 12 years old, 26 males). Demographic and behavioral information was assessed through standardized clinical tests as described in our recent works^9,27,34^ and reported in Table 1. Participants’ cognition was assessed with the Oxford Cognitive Screen (OCS),^35^ and language tests comprised the Nederlandse Benoem Test (NBT), i.e. Dutch naming test,^36^ and the ScreeLing test,^37^ which is aimed at assessing phonological, semantic and syntactic abilities. Although 12 PWA did not score below the cut-off scores in either language tests on the session day, they had a documented language impairment in the acute phase and were still attending speech-language therapy sessions at the time when the testing sessions took place.

**Table 1.** Demographic and behavioural data.

|  |  | HC (n = 24) | PWA (n = 40) | Statistics |
| --- | --- | --- | --- | --- |
| Demographics | Age (years old) | 72 ± 7 | 73 ± 11 | $U = 1285.5, P = 0.846^a$ |
| | Sex (male/female) | 16/8 | 26/14 | $\chi^2 = 0.019, P = 0.892^b$ |
| Cognition | **OCS total score**<br>Composite %: attention, memory and executive function<sup>35</sup> | 93.95 ± 4.62 | 79.56 ± 15.74 | $U = 569, P < 0.001^a$ |
| Language tests | **NBT total score**<br>Participants were asked to describe a series of images in one word each<sup>36</sup> | 271.30 ± 4.09 | 224.13 ± 57.72 | $U = 845.5, P < 0.001^a$ |
| | **ScreeLing total score**<br>Three parts with each four different tasks, covering semantics, phonology and syntax (El Hachoui et al., 2012). | 69.78 ± 2.49 | 62.17 ± 9.67 | $U = 563.5, P < 0.001^a$ |

<sup>a</sup>Unpaired Mann–Whitney U-test
<sup>b</sup>Chi-squared test

### 2.2. Experimental design

All experimental sessions took place in a soundproof room with a Faraday cage at the Department of Neurosciences, KU Leuven. EEG signals were recorded while participants listened to a 25-minute-long fairy tale titled “De Wilde Zwanen” (“The Wild Swans”) by Hans Christian Andersen, narrated by a female Flemish-native speaker. Throughout the listening, every five minutes a break was introduced and questions about the story to that point were asked to ensure participant’s attention, resulting in five 5-min story segments. The speech stimulus was presented binaurally through ER-3A insert phones (Etymotic Research Inc, IL, USA) using the software APEX.^38^ Participants were deemed hearing impaired if any hearing threshold was higher than 25 dB hearing loss on frequencies below 4 kHz. To ensure audibility in this case, stimulus intensity was set at 60 dB SPL plus half of the pure tone average (PTA) of the individual audiometric thresholds at 250, 500 and 1000 Hz. EEG signals were obtained using a Biosemi ActiveTwo (Amsterdam, Netherlands) cap with 64 Ag/AgCl electrodes distributed according to the 10-20 system^39^ as shown in Figure 1. Signals were recorded at a sampling rate of 8192 Hz.

**Figure 1.**
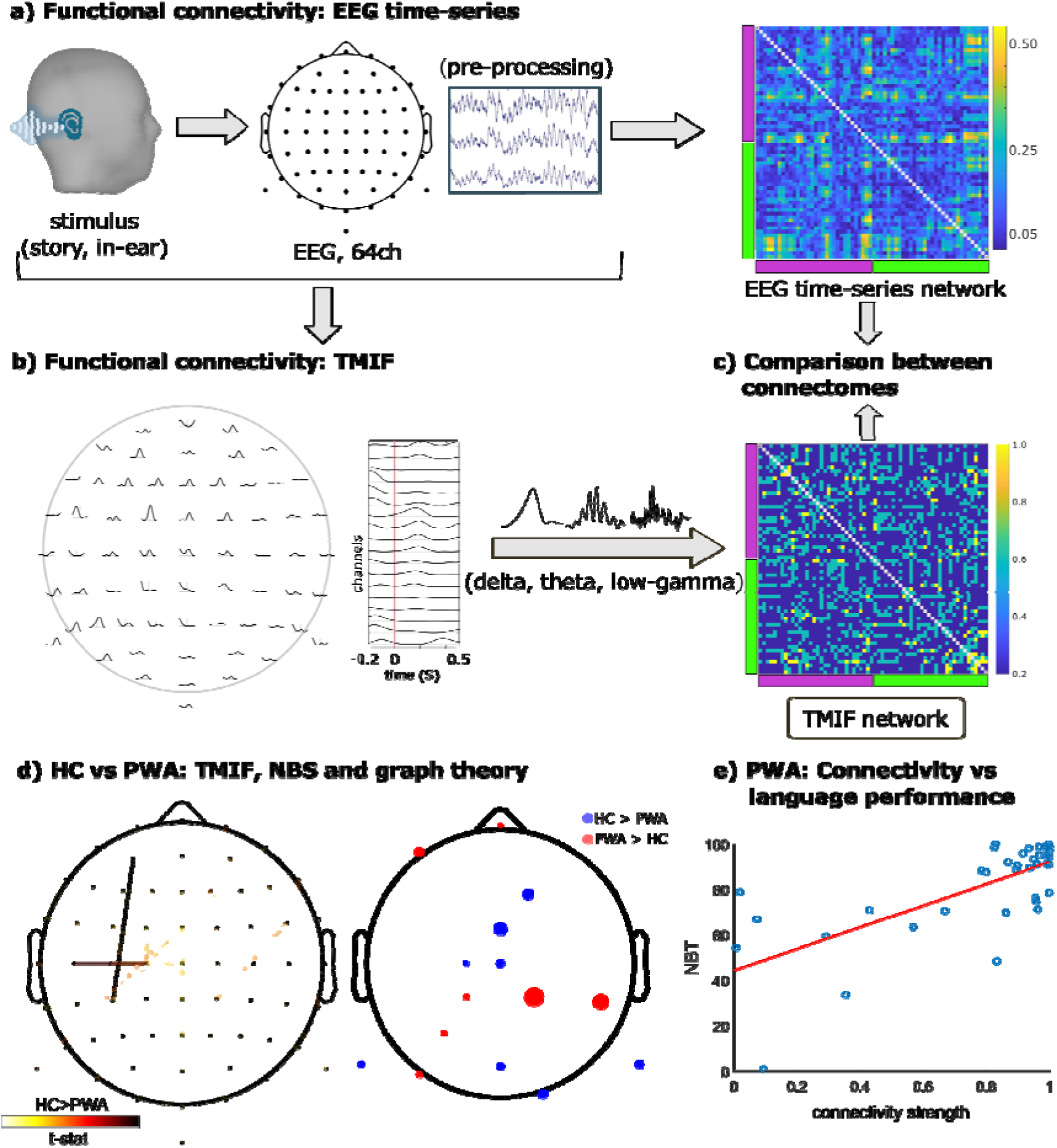
Methodological workflow. a) EEG is recorded during natural speech listening, and functional connectivity is extracted from the recorded signals. On the connectivity matrix, purple bar: left hemisphere; green bar: right hemisphere. b) Temporal mutual information functions (TMIFs) are computed for each EEG channel within delta, theta and low-gamma bands, and corresponding TMIF- connectomes are computed. c) Functional connectomes are compared between the two approaches. Magenta bar: left-hemisphere channels; green bar: right-hemisphere channels. d) TMIFs and functional connectomes obtained from TMIFs are compared between groups with the Network Based Statistics (NBS) and graph theory. On the second topography, blue: HC > PWA, red: PWA > HC. e) Correlation between TMIF-connectivity strength and language performance.

### 2.3. Speech envelope extraction

The envelope of the speech signal was extracted with a gammatone filter bank. The filter was implemented with 28 channels spaced by one equivalent rectangular bandwidth and center frequencies spanning between 50 Hz and 5000 Hz. To extract the envelopes at each sub-band, the absolute value of each sample was computed and raised to the power of 0.6. The speech envelope was yielded by averaging the envelopes across sub-bands. The obtained signal was downsampled to 512 Hz and filtered in the frequency ranges of interest. High- and lowpass least squares filters of order 2000 were implemented with a transition band of 10% below the highpass and 10% above the lowpass frequency, and compensation for group delay. The envelope was eventually normalized and further subsampled to 128 Hz.

### 2.4. EEG data pre-processing

Pre-processing of the EEG data was automated as implemented in the Automagic toolbox^40^ and is described in detail in our previous works.^9,27^ Briefly, signals were filtered to remove low-frequency (cut-off: 0.1 Hz) and line-noise, subsampled to 512 Hz, and cleaned from bad channels (average number of removed channels: 2 ± 2). Sporadic artifacts were time-interpolated, whilst systematic artifactual patterns were removed by means of independent component analysis (ICA) (average number of removed components: 26 ± 7). Before any further analysis, all signals were inspected visually, average-referenced and subsampled to 128 Hz.

### 2.5. Neural envelope tracking

As described in detail in our previous publication,^27^ we here also used the Gaussian copula mutual information (MI) analysis to assess neural tracking of natural speech.^41^ Compared to other methods, the Gaussian copula is reportedly unaffected by biases associated with the power spectrum of the signal and the stimulus and can effectively handle high-dimensional data.^26^ The copula was applied to the MI between the EEG signal and the stimulus, defined as:

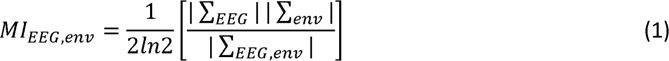

where the summation operators are the determinants of the covariance matrices of respectively EEG, envelope, and the joint variable. The temporal dynamics of MI were obtained by progressively shifting the EEG signal with respect to the speech envelope and computing the metric as defined in Equation (1) at each time-lag. The shift was performed within an integration window between −200 ms and 500 ms in order to capture stimulus-response latencies of interest.^26^ This process was performed for all EEG channels, and the obtained neural-tracking-metric consisted of the TMIF.^26,42^ To avoid artefactual effects due to discontinuity across recording segments (see Supplementary Fig. 2) and due to the fact that one participant fell asleep sporadically after the first 5-min recording, TMIF was only computed for the first 5-min segment, since such an amount was shown to already provide an estimation of neural tracking comparable to the one obtained from the whole EEG recording in both HC and PWA.^27^

### 2.6. Functional connectivity

Connectivity between the EEG time series and between the TMIFs across channels was measured with the weighted phase lag index (WPLI),^43^ defined as:

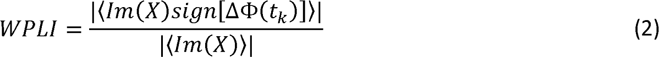

where *X* is the cross-spectrum between the signals, *Im(X)* is its imaginary part, Δϕ is the phase difference between the signals, t_k_ is the time step with k = 1, 2, …, *N* and *sign* is the signum function. WPLI computation was based on instantaneous phase differences measured through Hilbert transform, as implemented in the Brainstorm toolbox.^44^ Connectivity was measured for delta, theta and low-gamma band networks.

### 2.7. Graph theory

In the present work, we computed the same graph measures as we reported in our previous research.^9^ These include node strength, clustering coefficient, eccentricity, characteristic path length, small-worldness, and modularity. Functions to compute these metrics are included in the Brain Connectivity Toolbox (BCT) for MATLAB.^45^ Before computing the network measures, graphs were obtained by applying proportional thresholding on connectivity matrices to preserve between 3-40% of network density. This range reportedly reflects the underlying structural network properties, while allowing to take into account the dependence of the measures on the network size.^46–51^ Connectivity matrices were then normalized by their maximum weight,^52^ and graph metrics were computed at each network density. Further details on the graph theory metrics are reported in Supplementary materials of the publications by Mehraram and colleagues (2023), Rubinov & Sporns (2010) or Kaiser (2011).

### 2.8. Statistical analysis

#### EEG time series vs TMIF-connectomes

WPLI matrices were obtained from the first five minutes of the EEG signals and compared with the connectomes based on the TMIFs extracted from the same recording segment. We limited this analysis to the HC group and used the Network Based Statistics (NBS) toolbox^53^ to define a generalized linear model (GLM) with the methodological approach (EEG time series vs TMIF) as within-subject factor. The primary threshold of the NBS was set to 3, and a t-test for each direction was performed with 5000 permutations across the two methods (*P* < 0.025). If any significant network component was obtained, the distribution of the altered connectivity patterns across participants was inferred by generating the corresponding graph with the support of functions included in the EEGLAB toolbox^54^ and by visualizing the average connectivity within the NBS component (WPLI_NBS_) for both the EEG time series and TMIF.

#### TMIF-connectome: PWA vs HC

TMIFs and functional networks obtained from TMIF signals were compared between HC and PWA. Comparison between TMIFs at all frequency bands was performed with non-parametric spatiotemporal cluster-based permutation tests with 10000 permutations (*P* < 0.01)^55^ as implemented in the Eelbrain toolbox for Python.^56^ Comparison between connectomes was performed implementing an F-test with the NBS. Within the latter, primary threshold was set to 3, and significant network components were detected by performing 5000 permutations (*P* < 0.05). Direction of significance of any significant component was inferred by visualizing the distribution of WPLI_NBS_ across groups.

#### Graph theory: PWA vs. HC

Graph measures were compared between groups at each network density with Mann-Whitney U-tests (*P* < 0.05). Nodal measures, i.e., node strength and clustering coefficient, were locally tested for each node (64 electrodes, uncorrected for multiple tests) as well as globally on average across nodes. Graph theory analysis was only performed within frequency ranges which yielded significant results from the between-group NBS test described in the previous paragraph.

#### TMIF-connectivity vs language performance

We used the NBS also to test whether the TMIF-connectome of PWA correlates with either the score of the ScreeLing test or the NBT. For the correlation test, we tested a range of primary thresholds between 1-20, and significant results are reported in the next section. Results were also controlled for lesion size and age.

## 3. Results

### 3.1. Demographic data

Age range was not significantly different between groups (Mann-Whitney U-test, *U* = 1285.5, *P* = 0.846), and the ratio of males versus females was not different (Chi-squared test*, χ*^2^ = 0.019, *P* = 0.892). The HC group performed better in all behavioral language and cognitive tests (OCS: *U* = 569, *P* < 0.001; NBT: *U* = 845.5, *P* < 0.001; ScreeLing: *U* = 563.5, *P* < 0.001). Statistics on demographic data and behavioral test scores are reported in Table 1.

### 3.2. Functional connectome: TMIF vs EEG time series

We used the NBS to investigate the added value of analyzing the functional connectome associated with the TMIF versus EEG time series when investigating natural speech processing in the brain. The NBS yielded significant results only when testing in the TMIF > EEG_time series_ direction, i.e. WPLI in the connectome associated with the TMIFs is higher than in the one associated with EEG time series (delta: t ≤ 4.8, *P* = 0.02; theta: t ≤ 4.6, *P* = 0.005; low-gamma: t ≤ 7.8, *P* < 0.001) (Figure 2). Paired tests produced a stronger network component derived from the TMIFs, i.e. a region-specific emphasized synchronization between speech-related brain responses, as compared with the EEG time series. In the delta band, the cluster was centralized over the right temporal and parietal region, and also included connections between occipital, frontal and widespread left-hemispheric channels. On the contrary, the theta-band network shows higher centralization over the left hemisphere for the TMIF-estimated connectome, including central and prefrontal scalp regions. The low-gamma TMIF-connectome shows stronger connectivity compared to the EEG-estimated network mainly over posterior-anterior paths. Topographies from one-sample t-tests are reported in Supplementary material (Supplementary Fig. 1).

**Figure 2.**
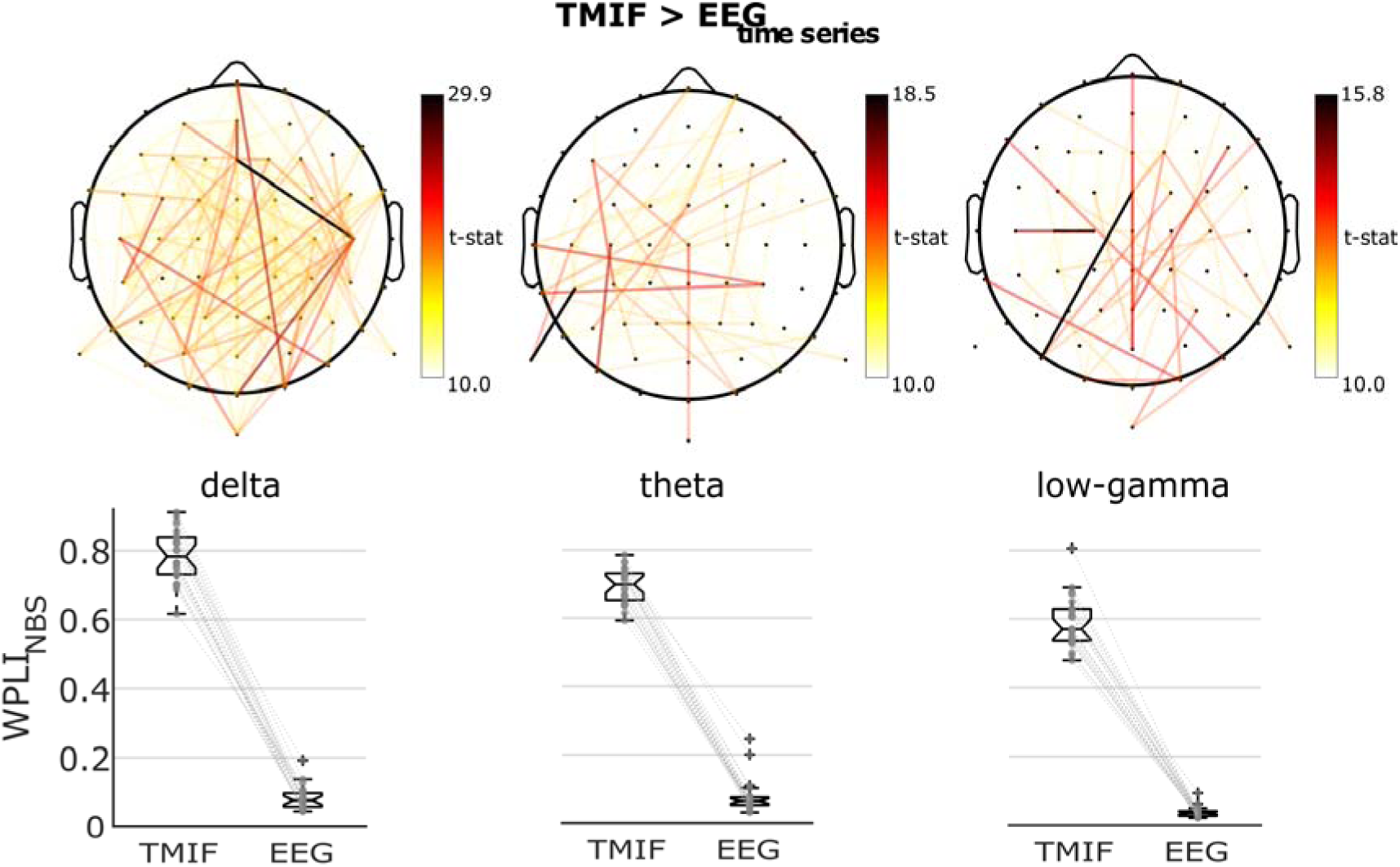
NBS outcome: TMIF vs EEG time series. The functional connectome associated with the neural envelope tracking exhibits a more strongly connected component when compared to the EEG time series-related network.

### 3.3. Between-group network comparison

The outcome of our analysis on the TMIFs is reported in Figure 3. Similar to the results from our previous work, PWA showed lower MI values compared to HC in the delta and theta band, whilst no difference emerged from the low-gamma band (Figure 3A). Cluster-based permutation tests yielded the strongest effect in the delta band within a spatial cluster with 15 channels between 89 – 300 ms after the stimulus onset (P = 0.004), whilst the theta band exhibited four clusters, respectively with 48 channels between −141 – 34 ms (P < 0.001), 47 channels between 34 – 120 ms (P = 0.001), 45 channels between 120 – 206 ms (P = 0.001), and 48 channels between 214 – 323 ms (P < 0.001). The NBS yielded significant results from the between-group comparison only within the low-gamma band (Figure 3B). Namely, low-gamma-network connectivity was weaker in PWA compared to HC, with widespread effect mostly marked over the left-hemisphere involving central and frontal scalp regions (F ≤ 18.9, P = 0.043) (topographies from one-sample t-tests for delta and theta bands are reported in Supplementary Fig. 1). Between-group differences also emerged from Graph Theory analysis (Figure 3C). On average, connectivity strength of the individual nodes was reduced in PWA only when network density was lower than 21%, with the effect mostly driven by central regions. Nevertheless, local differences in the opposite direction, i.e., PWA > HC, also emerged, namely over left parietal and temporal scalp regions. Global graph measures, i.e., average characteristic path length, small-worldness and modularity, were all higher in PWA, with the first two showing an effect only when network densities were respectively higher than 34% and 17% of the strongest connections. Results were deemed significant at *P* < 0.05 (uncorrected for multiple comparisons due to intrinsic interdependence of observations).

**Figure 3.**
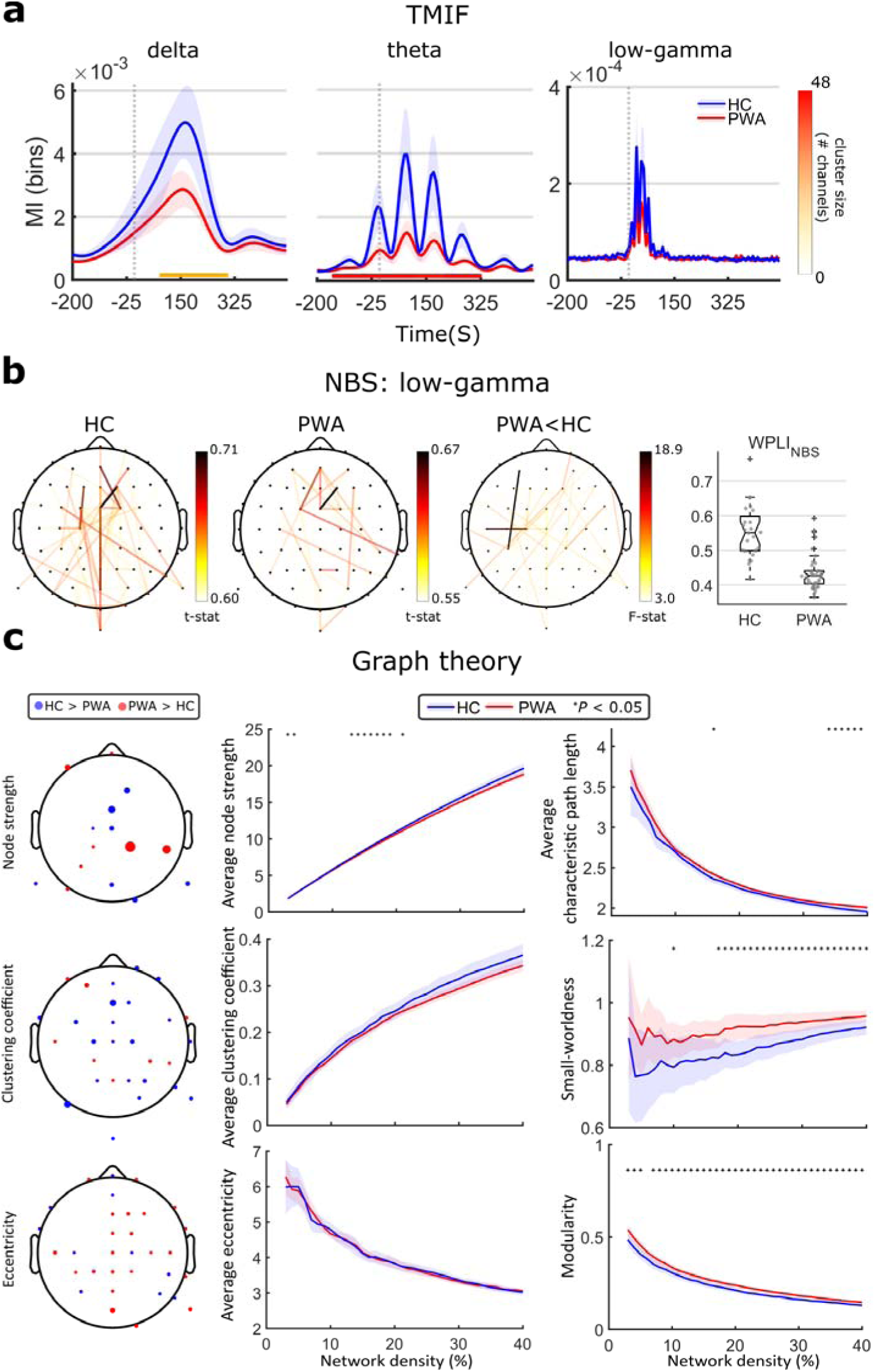
Comparison of neural tracking metrics between PWA and HC. a) TMIF-values were averaged across channels and compared between groups throughout the integration window. Time corresponding to a cluster associated with significant difference is highlighted with the faded bar on the bottom (with color reflecting cluster size as number of EEG channels). The dotted gray line corresponds to the stimulus onset. Shaded area corresponds to 95% confidence interval. b) Outcome of the NBS analysis. Topographies from left to right correspond respectively to one-sample t-test within HC group, one-sample t-test within PWA group, and F-test testing for PWA vs HC. A left-hemispheric network component is weakened in PWA compared to HC. c) Graph theory analysis. On average, PWA feature higher modularity and small-worldness. Shaded area corresponds to 95% confidence interval.

### 3.4. Connectivity strength vs language performance in PWA

We used the NBS to investigate whether there exists any subnetwork that correlates with the performance of language tests, namely the NBT and the ScreeLing test. A significant outcome only emerged in the delta band, consisting of positive correlation between one link over the left-prefrontal region and the NBT score, as shown in Figure 4 (F = 34.4, *P* = 0.026). Nevertheless, no significant result was found when lesion size and age were included in the GLM as nuisance covariates.

**Figure 4.**
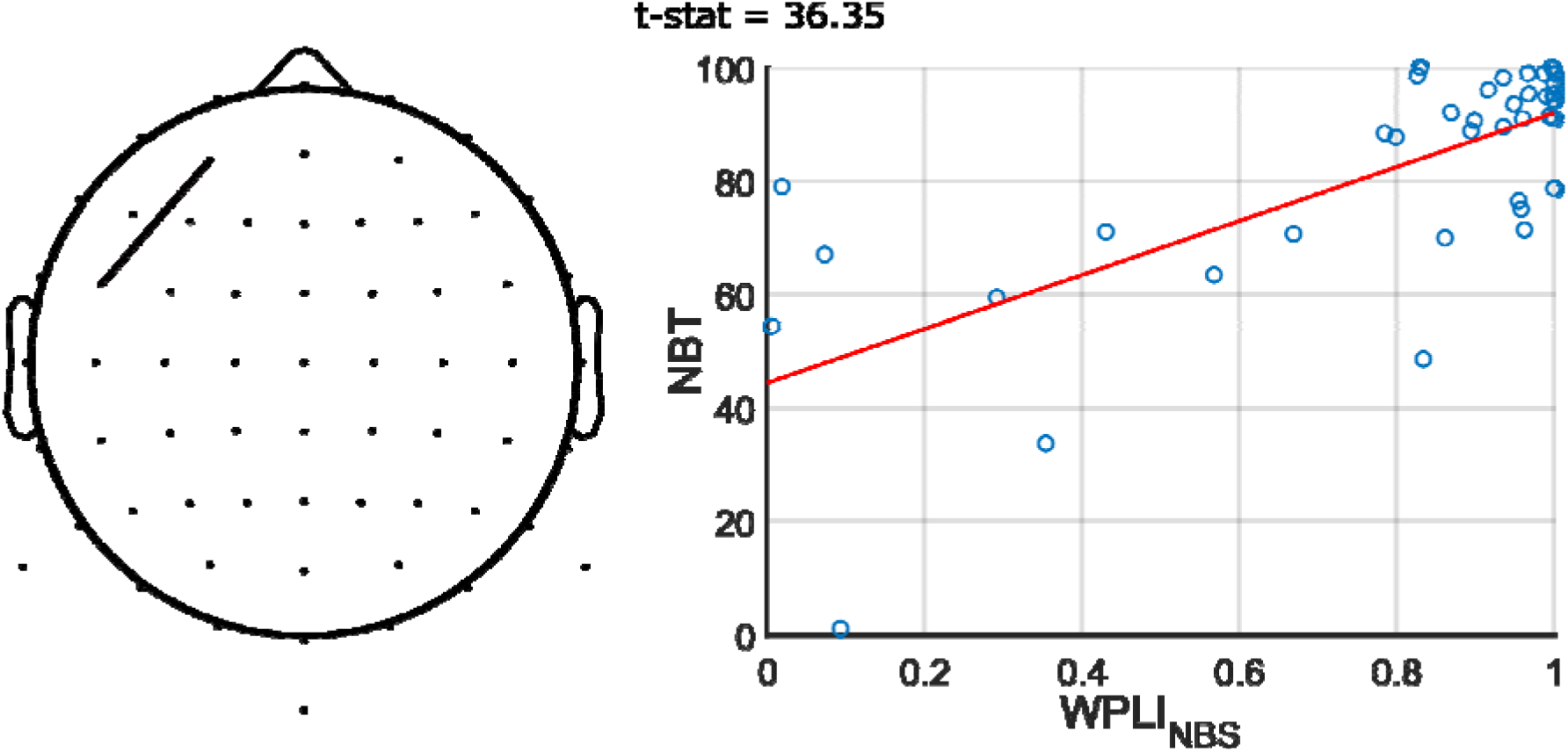
Outcome of NBS correlation tests with language performance. Only a positive correlation between delta-band connectivity and NBT percentage score was found.

## 4. Discussion

In the present study, we used EEG and we combined neural envelope tracking with connectomics to investigate natural speech processing in PWA. We first assessed the added value of the combined approach as compared to the analysis of the connectome solely based on EEG time series, and found a more prominent subnetwork in the TMIF-connectome. The latter was affected in PWA when compared with HC only in the low-gamma band, showing weaker connectivity and more segregated network distribution, whilst delta-band network connectivity was associated with impairment of language performance in PWA.

### 4.1. Connectivity patterns are more consistent in TMIFs compared to EEG time series

Using the NBS within the HC group, we implemented a GLM to assess whether functional networks obtained from TMIFs reveal significant region-specific patterns compared to the connectome associated with the EEG time series. Functional connectivity based on TRFs was already investigated in a recent EEG study investigating natural speech processing in healthy participants. The authors successfully demonstrated that the response to speech stimulus is synchronized across brain regions, and that such synchronization is organized in a community structure, hence defining a functional connectome. We here confirmed the suitability of such an approach, now based on TMIFs instead of TRFs, as opposed to the analysis of whole EEG time series when analyzing the brain response to natural speech.

In the delta band we found a subnetwork connected with the right temporal and parietal lobes statistically dependent on the signal from which the connectome is obtained, i.e. TMIF vs EEG time series, with TMIFs being associated with higher connectivity. As consistently reported in the literature on speech processing, EEG delta-band is commonly associated with acoustic modulations, such as prosody and rhythm, and sound articulation.^12,13,58,59^ As hypothesized in the Asymmetric Sampling in Time (AST) theory, these processes feature lateralization towards the right auditory cortex,^60–62^ and reflect into the neural envelope tracking. Our comparative analysis suggests that the emergence of these mechanisms from the EEG time series might be hindered due to other cognitive processes being captured at the same time, resulting in low signal-to-noise ratio (SNR). On the other hand, extraction of neural tracking-related signal, i.e. TMIF, allows to isolate the mechanisms of interest and obtain a signal solely reflecting the corresponding cortical activity, hence achieving a higher SNR. Higher connectivity in a subnetwork centralized over the right temporal lobe is in line with the AST hypothesis, and it favors connectome-analysis based on TMIF rather than EEG time series to aim for more interpretable outcomes when investigating natural speech processing.

A significant outcome was also yielded in the theta band, consisting of a widespread TMIF-network with stronger connectivity compared to the EEG-time-series connectome. According to the AST, processing at the syllable rate is associated with theta-band activity.^13,59,61^ Hence, our result is evidence of higher sensitivity of TMIF compared to EEG time series to also estimate the connectome associated with processing of this language feature.

The strongest effect was detected in the low-gamma band, where the TMIF-connectome exhibits stronger posterior-anterior connectivity compared to the EEG time-series network. The actual role of gamma-band activity is still debated among researchers.^63,64^ Coupled with theta activity, gamma oscillations are known to reflect inhibitory activity of interneurons associated with memory, attentional and learning mechanisms.^65–68^ By assessing the connectome associated with the brain response to speech, we aimed at limiting the gamma activity of interest to language-related processes, reportedly phonemic processing,^30,60,66^ and these are likely reflected into the obtained topography.

### 4.2. TMIF-connectome is weakened in PWA

Before analyzing functional networks, we compared TMIFs across groups as a validation of the results reported in our previous work.^27^ With a larger sample than in the previous study, we also obtained reduced neural tracking in PWA in delta and theta bands, confirming the robustness of the TMIF waveform as a biomarker for post-stroke aphasia. However, no difference emerged in the low-gamma band. For the latter, statistical power might be affected either by the sample size, recording length, channel selection, single-channel here vs multivariate TMIF computation in De Clercq et al.^27^ or the stricter threshold in the cluster-detection algorithm (*P* = 0.01 in this study against *P* = 0.05 in De Clercq et al.^27^).

We then measured connectivity between TMIF signals and compared the obtained connectomes between PWA and HC using the NBS. In contrast with the previous analysis where TMIF signals were compared, a between-group difference only emerged in the low-gamma band when TMIF connectomes are compared. We found a weakened widespread interhemispheric frontal-posterior subnetwork in PWA, with a strong effect over the left-prefrontal region. Reduced interhemispheric connectivity is a common consequence of stroke,^8^ and together with the fact that the main effect is located over the most frequent lesion site across participants (see the lesion overlap map in Supplementary Fig. 3), our finding may here reflect impairment of phonemic and phonetic processing in PWA.^30,60,66^ A difference in the functional connectome in the low-gamma band is in line with our previous study based on EEG time series, where we also reported reduced posterior-anterior connectivity.^9^ The fact that a difference in the connectome was not found in the theta band may suggest that the corresponding outcome in our previous study might be reflecting other language-related processes, such as cognitive or executive functions. Nevertheless, this remains speculative and should be further investigated in future studies.

We believe that these results further prove the additive value of connectomics approach on the analysis of neural tracking of continuous natural speech in post-stroke aphasia. A commonly pursued approach in the research on neural tracking consists of averaging the waveforms within certain regions of interest (ROIs) before performing any between- or within-group-comparison.^58,69,70^ However, this can result in attenuation of the average signal in PWA due to the misalignment of the waveforms compared to HC, hence leading to inaccurate neurophysiological interpretation. In fact, reduced connectivity between TMIFs reflects a lack of synchronization between stimulus-responses across channels. On the other hand, the NBS is a data-driven approach, and revealed localized significant alterations of connectivity in PWA, proving that neural-tracking pathological abnormalities are localized and emerge at a network level. Therefore, we believe that it would be valuable for future neural-tracking studies to be complemented with data-driven network analysis for effective assessment of response-related brain regions. This strategy may disentangle methodological obliquities, let the research efficiently focus on regions involved in the process of interest, and provide further topographical information associated with the neural response to speech.

### 4.3. The functional network tends towards a small-world organization in PWA

To further disentangle alterations in the functional connectome in PWA, we performed graph theory analysis. Globally, TMIF-networks in PWA showed reduced network integration as compared to HC, but only when weaker connections, i.e. low synchronization between TMIFs across regions, were preserved in the definition of the graph via thresholding. This finding may reflect a preservation of network integration within the strongest connections of the network, potentially as a compensatory mechanism for the affected weaker links. Despite a weakened connectivity pattern in PWA being detected with the NBS as also reflected in the average node strength, local increases in functional connectivity in PWA also emerged, namely over the right temporal and central scalp regions. This latter finding resonates with existing evidence in fMRI studies reporting compensatory mechanism driven by preserved regions within the contra-lesional hemisphere, associated with recovery towards the chronic stage.^71–74^ The fact that this feature was found in connectomes associated with neural envelope tracking of speech reinforces the link between physiological compensatory mechanism and language impairment, excluding a sole relation to stroke-related anatomical alterations. Local increase in connectivity strength in PWA, although significant, may not be defining a connected cluster to be detected with the NBS.

We also found higher small-worldness in PWA compared to HC. A small-world network features a high level of clustering whilst integration is similar to a random network of the same size.^75–77^ Small-worldness is normally associated with network complexity and efficiency,^48,78,79^ and enhanced small-worldness in the TMIF-connectome in PWA might appear paradoxical. This might be interpreted in the context of compensatory mechanisms associated with aphasia, as discussed above, where increased node strength within the right temporal and parietal regions might be driving a global reorganization of the network favoring higher functional efficiency. Interestingly, higher small-worldness was also reported for other pathological conditions, including dementia with Lewy bodies,^46,49^ sleep deprivation,^80^ and post-traumatic stress disorder associated with traumatic brain injury.^81^ It is likely that these conditions share local increase in connectivity and prevalence of short connections, that we also reported in PWA compared to HC. This alteration reflects a reorganization of the network towards a more regular topology,^75^ and results in higher small-worldness. Small-worldness together with higher modularity in PWA also suggest increased network segregation compared to HC, reflecting an overall damage of the functional connectome.

### 4.4. Delta-band connectivity is associated with aphasia severity

Using the NBS we found a correlation between delta-band connectivity and the performance of NBT in PWA. Specifically, weaker connectivity over the left prefrontal region is associated with worse performance in the Dutch naming test. Association between delta-band connectivity and language performance agrees with the results reported in our previous study,^9^ and further favors the relevance of delta oscillation for the assessment of the severity of the pathology. As discussed in the previous paragraph, a network pattern in the delta band is likely to reflect neurophysiological mechanisms associated with perception of rhythm, acoustic modulation and lexical processing.^12,13,58,59,82^ Hence, affected neural-tracking connectivity may relate to disruption of rhythmic, acoustic and lexical processing in post-stroke aphasia as emerging in a naming task. However, the sample distribution features a ceiling effect, which might have attenuated the statistical power of this analysis and limited the effect to only one connection. Together with non-significant result when also testing for correlation with the ScreeLing test, this outcome strengthens the relevance of ensuring enough within-group behavioral variability at the recruitment stage. Interestingly, despite the intrinsic variability of anatomical properties of the lesion(s) as also emerges from the lesion overlap map reported in Supplementary Fig. 3), the functional connection associated with language performance in PWA is located over the most frequently lesioned area. However, the correlation is not significant when lesion size is included as nuisance covariate. As discussed in the next section of this paper, this might potentially reveal a statistical confound related to anatomical properties of the lesion, which should be addressed with appropriate lesion modelling.

### 4.5. Limitations

Measurement of neural tracking requires choosing a specific speech-stimulus feature. Hence, a single TMIF does not allow assessment of the brain response to the overall speech stimulus. This may result in omitting relevant biomarkers not associated with the feature of interest. Nevertheless, as we discussed in the Introduction section, the envelope of speech reportedly contains most of the energy of the stimulus signal and suffices for an optimal perception of speech.^14–19^

The electrical signals recorded with EEG correspond to spatial linear combinations of cortical activation. The latter are influenced by conductivity of anatomical layers adjacent to the cortical tissue.^83^ In the case of a stroke this phenomenon is likely enhanced due to the associated cortical lesion,^84,85^ potentially introducing unpredictable biases in the statistical analysis and, in this paper, lower SNR in the computation of TMIF connectomes in PWA. Nevertheless, aphasia features a high level of pathological variability,^86^ and alteration in volume conductivity is expected to introduce a confounding factor, resulting in weaker statistical power. With the NBS we detected a significantly affected subnetwork in PWA and demonstrated the consistency of language-related functional alteration despite variability in stroke characteristics. Future studies should implement source-level analysis with accurate modelling of volume conductivity of brain tissues in order to address lesion-related signal distortions.^87,88^

Aphasia also involves impairment of cognitive functions. This raises the concern whether this might be driving the detected between-group differences, questioning the interpretability of our results. Due to the intrinsic comorbidity of post-stroke aphasia, disentangling this aspect remains challenging, although we believe that our natural-speech-based experimental paradigm favored the emergence of speech-processing-related activity in the EEG measurement, thus attenuating cognition-related confounding factors.

Another limitation lies in the chosen connectivity metric, i.e. WPLI. In fact, this phase-based measure lacks information on directionality, hence interpretation of the results remains limited. In addition, almost-zero-lag connections are omitted, whilst they may contain relevant topological information. Alternative metrics, such as amplitude correlation^11^ or Granger causality,^89^ may be considered to obtain complementary additional insights into the functional connectome in PWA.

A component of arbitrariness is introduced with the choice of a primary threshold in the NBS.^53^ In the present work, we selected permissive values and made statistical interpretations based on t- or F-statistics of the connections within the obtained network components.

The comparisons of connectomes by means of the NBS was controlled for the family wise error rate. However, correction for multiple tests for local graph theory measures remains challenging, due to the high number of tests, interdependence of the recorded cortical activity at different EEG channels and the exploratory nature of our study.

## 5. Conclusion

We investigated the functional connectome associated with neural tracking of natural speech in PWA. We reported reduced connectivity between cortical stimulus-responses in PWA compared to HC, as well as altered network topology. Whilst exhibiting reduced functional connectivity, the connectome in PWA also tends towards a more segregated architecture likely reflecting a damage of physiological natural speech processing. We also showed a correlation between TMIF-connectivity and language performance in PWA, demonstrating the potential of connectomics-based approaches for the assessment of severity of post-stroke aphasia. Overall, our study shows the added value of connectomics to the analysis of natural speech processing in PWA when performed in a combined fashion with neural envelope tracking. Future research should investigate whether such an approach may also effectively define informative biomarkers for the assessment of aphasia at the acute stroke.

## Data Availability

Raw EEG data can be made available upon reasonable request to the authors by qualified researchers and if the GDPR-related conditions are met

## 7 Acknowledgments

The authors express their gratitude to the participants for their valuable contribution to this research work. We also thank Dr. Klara Schevenels (ORCID: 0000-0002-8474-3690) for her assistance in the recruitment process, as well as the individuals that were involved in the data collection. This research was funded by the Research Foundation Flanders (FWO; PhD grant P.D.C. 1S40122N, grant G0D8520N), the Luxembourg National Research Fund (FNR; AFR-PhD grant J.K. 13513810) and the European Research Council (ERC) under the European Union’s Horizon 2020 research and innovation programme (T.F., No. 637424).

## 8 Author contributions

R.M. implemented the analyses and produced the present manuscript with contributions from all authors. R.M., J.K., P.D.C., M.V. and T.F. conceived the study and designed the experiment. J.K., P.D.C. and R.M. collected the data.

## 9. Data availability

The ethical approval of the present study does not permit public archiving of raw neuroimaging data, but raw EEG data can be made available upon reasonable request to the corresponding author (R.M.) by qualified researchers and if the GDPR-related conditions are met.

## 10. Additional information

The authors declare no competing interests.

## References

1. Gialanella B, Bertolinelli M, Lissi M, Prometti P. Predicting outcome after stroke: the role of aphasia. https://doi.org/103109/096382882010488712. 2010;33(2):122–129. doi:10.3109/09638288.2010.488712

2. Flowers HL, Skoretz SA, Silver FL, et al. Poststroke Aphasia Frequency, Recovery, and Outcomes: A Systematic Review and Meta-Analysis. Arch Phys Med Rehabil. 2016;97(12):2188–2201.e8. 10.1016/j.apmr.2016.03.006

3. Rohde A, Worrall L, Godecke E, O’Halloran R, Farrell A, Massey M. Diagnosis of aphasia in stroke populations: A systematic review of language tests. PLoS One. 2018;13(3):e0194143. doi:10.1371/JOURNAL.PONE.0194143

4. Meng Q, Zeng Q, Xie Q, et al. Flexible lower limb exoskeleton systems: A review. NeuroRehabilitation. 2022;50:367–390. doi:10.3233/NRE-210300

5. Li R, Mukadam N, Kiran S. Functional MRI evidence for reorganization of language networks after stroke. Handb Clin Neurol. 2022;185:131–150. doi:10.1016/B978-0-12-823384-9.00007-4

6. Hartwigsen G, Saur D. Neuroimaging of stroke recovery from aphasia – Insights into plasticity of the human language network. Neuroimage. 2019;190(August 2017):14–31. doi:10.1016/j.neuroimage.2017.11.056

7. Saur D, Lange R, Baumgaertner A, et al. Dynamics of language reorganization after stroke. Brain. 2006;129(6):1371–1384. doi:10.1093/brain/awl090

8. Siegel JS, Ramsey LE, Snyder AZ, et al. Disruptions of network connectivity predict impairment in multiple behavioral domains after stroke. Proc Natl Acad Sci U S A. 2016;113(30):E4367–E4376. doi:10.1073/pnas.1521083113

9. Mehraram R, Kries J, De Clercq P, Vandermosten M, Francart T. EEG reveals brain network alterations in chronic aphasia during natural speech listening. bioRxiv. Published online March 10, 2023:2023.03.10.532034. doi:10.1101/2023.03.10.532034

10. Caliandro P, Vecchio F, Miraglia F, et al. Small-World Characteristics of Cortical Connectivity Changes in Acute Stroke. Neurorehabil Neural Repair. 2017;31(1):81–94. doi:10.1177/1545968316662525

11. Shah-Basak P, Sivaratnam G, Teti S, et al. Electrophysiological connectivity markers of preserved language functions in post-stroke aphasia. Neuroimage Clin. 2022;34(November 2021):103036. doi:10.1016/j.nicl.2022.103036

12. Gross J, Hoogenboom N, Thut G, et al. Speech rhythms and multiplexed oscillatory sensory coding in the human brain. PLoS Biol. 2013;11(12). doi:10.1371/JOURNAL.PBIO.1001752

13. Di Liberto GM, O’Sullivan JA, Lalor EC. Low-frequency cortical entrainment to speech reflects phoneme-level processing. Current Biology. 2015;25(19):2457–2465. doi:10.1016/j.cub.2015.08.030

14. Peelle JE, Davis MH. Neural oscillations carry speech rhythm through to comprehension. Front Psychol. 2012;3(SEP):27077. doi:10.3389/FPSYG.2012.00320/BIBTEX

15. Shannon R V., Zeng FG, Kamath V, Wygonski J, Ekelid M. Speech Recognition with Primarily Temporal Cues. Science (1979). 1995;270(5234):303–304. doi:10.1126/SCIENCE.270.5234.303

16. Ding N, Simon JZ. Adaptive Temporal Encoding Leads to a Background-Insensitive Cortical Representation of Speech. Journal of Neuroscience. 2013;33(13):5728–5735. doi:10.1523/JNEUROSCI.5297-12.2013

17. Etard O, Reichenbach T. Neural Speech Tracking in the Theta and in the Delta Frequency Band Differentially Encode Clarity and Comprehension of Speech in Noise. Journal of Neuroscience. 2019;39(29):5750–5759. doi:10.1523/JNEUROSCI.1828-18.2019

18. Vanthornhout J, Decruy L, Wouters J, Simon JZ, Francart T. Speech Intelligibility Predicted from Neural Entrainment of the Speech Envelope. JARO - Journal of the Association for Research in Otolaryngology. 2018;19(2):181–191. doi:10.1007/S10162-018-0654-Z/FIGURES/6

19. Gillis M, Van Canneyt J, Francart T, Vanthornhout J. Neural tracking as a diagnostic tool to assess the auditory pathway. Hear Res. 2022;426. doi:10.1016/J.HEARES.2022.108607

20. Zhou D, Zhang G, Dang J, Unoki M, Liu X. Detection of Brain Network Communities During Natural Speech Comprehension From Functionally Aligned EEG Sources. Front Comput Neurosci. 2022;16:919215. doi:10.3389/FNCOM.2022.919215/BIBTEX

21. Dial HR, Gnanateja GN, Tessmer RS, Gorno-Tempini ML, Chandrasekaran B, Henry ML. Cortical Tracking of the Speech Envelope in Logopenic Variant Primary Progressive Aphasia. Front Hum Neurosci. 2021;14. doi:10.3389/FNHUM.2020.597694

22. Di Liberto GM, Peter V, Kalashnikova M, Goswami U, Burnham D, Lalor EC. Atypical cortical entrainment to speech in the right hemisphere underpins phonemic deficits in dyslexia. Neuroimage. 2018;175(November 2017):70–79. doi:10.1016/j.neuroimage.2018.03.072

23. Lizarazu M, Lallier M, Bourguignon M, Carreiras M, Molinaro N. Impaired neural response to speech edges in dyslexia. Cortex. 2021;135:207–218. doi:10.1016/J.CORTEX.2020.09.033

24. Mandke K, Flanagan S, Macfarlane A, et al. Neural sampling of the speech signal at different timescales by children with dyslexia. Neuroimage. 2022;253:119077. doi:10.1016/J.NEUROIMAGE.2022.119077

25. Kries J, Clercq P De, Gillis M, et al. Exploring neural tracking of acoustic and linguistic speech representations in individuals with post-stroke aphasia. bioRxiv. Published online March 2, 2023:2023.03.01.530707. doi:10.1101/2023.03.01.530707

26. De Clercq P, Vanthornhout J, Vandermosten M, Francart T. Beyond linear neural envelope tracking: a mutual information approach. J Neural Eng. 2023;20(2):026007. doi:10.1088/1741-2552/ACBE1D

27. De Clercq P, Kries J, Mehraram R, Vanthornhout J, Francart T, Vandermosten M. Detecting post-stroke aphasia using EEG-based neural envelope tracking of natural speech. medRxiv. Published online March 17, 2023:2023.03.14.23287194. doi:10.1101/2023.03.14.23287194

28. Giraud AL, Poeppel D. Cortical oscillations and speech processing: emerging computational principles and operations. Nature Neuroscience 2012 15:4. 2012;15(4):511–517. doi:10.1038/nn.3063

29. Ding N, Melloni L, Zhang H, Tian X, Poeppel D. Cortical tracking of hierarchical linguistic structures in connected speech. Nature Neuroscience 2016 19:1. 2015;19(1):158–164. doi:10.1038/nn.4186

30. Hyafil A, Giraud AL, Fontolan L, Gutkin B. Neural Cross-Frequency Coupling: Connecting Architectures, Mechanisms, and Functions. Trends Neurosci. 2015;38(11):725–740. doi:10.1016/J.TINS.2015.09.001

31. Flamand-Roze C, Falissard B, Roze E, et al. Validation of a new language screening tool for patients with acute stroke: the Language Screening Test (LAST). Stroke. 2011;42(5):1224–1229.

32. Schevenels K, Michiels L, Lemmens R, de Smedt B, Zink I, Vandermosten M. The role of the hippocampus in statistical learning and language recovery in persons with post stroke aphasia. Neuroimage Clin. 2022;36. doi:10.1016/J.NICL.2022.103243

33. Schevenels K, Price CJ, Zink I, de Smedt B, Vandermosten M. A Review on Treatment-Related Brain Changes in Aphasia. Neurobiology of Language. 2020;1(4):402–433. doi:10.1162/nol_a_00019

34. Kries J, De Clercq P, Lemmens R, Francart T, Vandermosten M. Tuning in on auditory details is difficult: Individuals with aphasia show impaired acoustic and phonemic processing. bioRxiv. Published online January 1, 2022:2022.12.14.520503. doi:10.1101/2022.12.14.520503

35. Demeyere N, Riddoch MJ, Slavkova ED, Bickerton WL, Humphreys GW. The Oxford Cognitive Screen (OCS): validation of a stroke-specific short cognitive screening tool. Psychol Assess. 2015;27(3):883–894. doi:10.1037/PAS0000082

36. van Ewijk L, Dijkhuis L, Hofs-van Kats M, Hendrickx-Jessurun M, Wijngaarden M, de Hilster C. Nederlandse Benoem Test. Published online 2018.

37. el Hachioui H, van de Sandt-Koenderman MWME, Dippel DWJ, Koudstaal PJ, Visch-Brink EG. The ScreeLing: occurrence of linguistic deficits in acute aphasia post-stroke. J Rehabil Med. 2012;44(5):429–435. doi:10.2340/16501977-0955

38. Francart T, van Wieringen A, Wouters J. APEX 3: a multi-purpose test platform for auditory psychophysical experiments. J Neurosci Methods. 2008;172(2):283–293. 10.1016/j.jneumeth.2008.04.020

39. Oostenveld R, Praamstra P. The five percent electrode system for high-resolution EEG and ERP measurements. Clinical Neurophysiology. 2001;112(4):713–719.

40. Pedroni A, Bahreini A, Langer N. Automagic: Standardized preprocessing of big EEG data. Neuroimage. 2019;200(June):460–473. doi:10.1016/j.neuroimage.2019.06.046

41. Ince RAA, Giordano BL, Kayser C, Rousselet GA, Gross J, Schyns PG. A statistical framework for neuroimaging data analysis based on mutual information estimated via a gaussian copula. Hum Brain Mapp. 2017;38(3):1541–1573. doi:10.1002/HBM.23471

42. Zan P, Presacco A, Anderson S, Simon JZ. Exaggerated cortical representation of speech in older listeners: mutual information analysis. J Neurophysiol. 2020;124(4):1152–1164. doi:10.1152/JN.00002.2020

43. Vinck M, Oostenveld R, van Wingerden M, Battaglia F, Pennartz CMA. An improved index of phase-synchronization for electrophysiological data in the presence of volume-conduction, noise and sample-size bias. Neuroimage. 2011;55(4):1548–1565. doi:10.1016/j.neuroimage.2011.01.055

44. Tadel F, Baillet S, Mosher JC, Pantazis D, Leahy RM. Brainstorm: a user-friendly application for MEG/EEG analysis. Comput Intell Neurosci. 2011;2011:879716. doi:10.1155/2011/879716

45. Rubinov M, Sporns O. Complex network measures of brain connectivity: Uses and interpretations. Neuroimage. 2010;52(3):1059–1069. 10.1016/j.neuroimage.2009.10.003

46. Mehraram R, Kaiser M, Cromarty R, et al. Weighted network measures reveal differences between dementia types: An EEG study. Hum Brain Mapp. 2020;41(6):1573–1590. doi:10.1002/hbm.24896

47. Bohr IJ, Kenny E, Blamire A, et al. Resting-state functional connectivity in late-life depression: higher global connectivity and more long distance connections. Front Psychiatry. 2013;3:116. doi:10.3389/fpsyt.2012.00116

48. Kaiser M. A tutorial in connectome analysis: Topological and spatial features of brain networks. Neuroimage. 2011;57(3):892–907. 10.1016/j.neuroimage.2011.05.025

49. Peraza LR, Taylor JP, Kaiser M. Divergent brain functional network alterations in dementia with Lewy bodies and Alzheimer’s disease. Neurobiol Aging. 2015;36(9):2458–2467. doi:10.1016/j.neurobiolaging.2015.05.015

50. Langer N, Pedroni A, Jancke L. The problem of thresholding in small-world network analysis. PLoS One. 2013;8(1):e53199. doi:10.1371/journal.pone.0053199

51. Rubinov M, Knock SA, Stam CJ, et al. Small-world properties of nonlinear brain activity in schizophrenia. Hum Brain Mapp. 2009;30(2):403–416. doi:10.1002/hbm.20517

52. Onnela JP, Saramäki J, Kertész J, Kaski K. Intensity and coherence of motifs in weighted complex networks. Phys Rev E. 2005;71(6):65103.

53. Zalesky A, Fornito A, Bullmore ET. Network-based statistic: identifying differences in brain networks. Neuroimage. 2010;53(4):1197–1207. doi:10.1016/j.neuroimage.2010.06.041

54. Delorme A, Makeig S. EEGLAB: an open source toolbox for analysis of single-trial EEG dynamics including independent component analysis. J Neurosci Methods. 2004;134(1):9–21. doi:10.1016/j.jneumeth.2003.10.009

55. Maris E, Oostenveld R. Nonparametric statistical testing of EEG- and MEG-data. J Neurosci Methods. 2007;164(1):177–190. doi:10.1016/J.JNEUMETH.2007.03.024

56. Brodbeck C, Das P, Brooks TL, Reddigari S, jpkulasingham. christianbrodbeck/Eelbrain: 0.39. Published online May 2023. doi:10.5281/zenodo.7951251

57. Zhu Y, Liu J, Ristaniemi T, Cong F. Distinct Patterns of Functional Connectivity During the Comprehension of Natural, Narrative Speech. https://doi.org/101142/S0129065720500070. 2020;30(3). doi:10.1142/S0129065720500070

58. Boucher VJ, Gilbert AC, Jemel B. The Role of Low-frequency Neural Oscillations in Speech Processing: Revisiting Delta Entrainment. Published online 2019. doi:10.1162/jocn_a_01410

59. Ding N, Simon JZ. Cortical entrainment to continuous speech: Functional roles and interpretations. Front Hum Neurosci. 2014;8(MAY):1–7. doi:10.3389/fnhum.2014.00311

60. Giraud AL, Kleinschmidt A, Poeppel D, Lund TE, Frackowiak RSJ, Laufs H. Endogenous Cortical Rhythms Determine Cerebral Specialization for Speech Perception and Production. Neuron. 2007;56(6):1127–1134. doi:10.1016/j.neuron.2007.09.038

61. Poeppel D. The analysis of speech in different temporal integration windows: cerebral lateralization as ‘asymmetric sampling in time.’ Speech Commun. 2003;41(1):245–255. doi:10.1016/S0167-6393(02)00107-3

62. Belin P, Fecteau S, Bédard C. Thinking the voice: neural correlates of voice perception. Trends Cogn Sci. 2004;8(3):129–135. doi:10.1016/J.TICS.2004.01.008

63. Buzsáki G, Wang XJ. Mechanisms of gamma oscillations. Annu Rev Neurosci. 2012;35:203–225.

64. Yuval-Greenberg S, Tomer O, Keren AS, Nelken I, Deouell LY. Transient induced gamma-band response in EEG as a manifestation of miniature saccades. Neuron. 2008;58(3):429–441. doi:10.1016/J.NEURON.2008.03.027

65. Belluscio MA, Mizuseki K, Schmidt R, Kempter R, Buzsáki G. Cross-Frequency Phase–Phase Coupling between Theta and Gamma Oscillations in the Hippocampus. The Journal of Neuroscience. 2012;32(2):423–435. doi:10.1523/jneurosci.4122-11.2012

66. Bartos M, Vida I, Jonas P. Synaptic mechanisms of synchronized gamma oscillations in inhibitory interneuron networks. Nat Rev Neurosci. 2007;8(1):45–56. doi:10.1038/nrn2044

67. Colgin LL, Denninger T, Fyhn M, et al. Frequency of gamma oscillations routes flow of information in the hippocampus. Nature. 2009;462(7271):353–357. doi:10.1038/nature08573

68. Kay LM. Two species of gamma oscillations in the olfactory bulb: dependence on behavioral state and synaptic interactions. J Integr Neurosci. 2003;2(1):31–44. doi:10.1142/S0219635203000196

69. Lesenfants D, Vanthornhout J, Verschueren E, Decruy L, Francart T. Predicting individual speech intelligibility from the cortical tracking of acoustic- and phonetic-level speech representations. Hear Res. 2019;380:1–9. doi:10.1016/J.HEARES.2019.05.006

70. Gillis M, Kries J, Vandermosten M, Francart T. Neural tracking of linguistic and acoustic speech representations decreases with advancing age. Neuroimage. 2023;267:119841. doi:10.1016/J.NEUROIMAGE.2022.119841

71. Cao Y, Vikingstad EM, George KP, Johnson AF, Welch KMA. Cortical Language Activation in Stroke Patients Recovering From Aphasia With Functional MRI. Stroke. 1999;30(11):2331–2340. doi:10.1161/01.STR.30.11.2331

72. Heiss W d, Kessler J, Thiel A, Ghaemi M, Karbe H, W-d H. Differential Capacity of Left and Right Hemispheric Areas for Compensation of Poststroke Aphasia. Published online 1999. doi:10.1002/1531-8249

73. Sims JA, Kapse K, Glynn P, Sandberg C, Tripodis Y, Kiran S. The relationships between the amount of spared tissue, percent signal change, and accuracy in semantic processing in aphasia. Neuropsychologia. 2016;84:113–126. doi:10.1016/j.neuropsychologia.2015.10.019

74. Turkeltaub PE, Messing S, Norise C, Hamilton RH. Are networks for residual language function and recovery consistent across aphasic patients? Neurology. 2011;76(20):1726–1734. doi:10.1212/WNL.0B013E31821A44C1

75. Telesford QK, Joyce KE, Hayasaka S, Burdette JH, Laurienti PJ. The ubiquity of small-world networks. Brain Connect. 2011;1(5):367–375. doi:10.1089/brain.2011.0038

76. Sporns O, Zwi JD. The small world of the cerebral cortex. Neuroinformatics. 2004;2(2):145–162.

77. Watts DJ, Strogatz SH. Collective dynamics of ‘small-world’networks. Nature. 1998;393(6684):440.

78. Bullmore E, Sporns O. Complex brain networks: graph theoretical analysis of structural and functional systems. Nat Rev Neurosci. 2009;10(3):186–198. doi:10.1038/nrn2575

79. Onoda K, Yamaguchi S. Small-worldness and modularity of the resting-state functional brain network decrease with aging. Neurosci Lett. 2013;556:104–108. 10.1016/j.neulet.2013.10.023

80. Liu H, Li H, Wang Y, Lei X. Enhanced brain small-worldness after sleep deprivation: a compensatory effect. J Sleep Res. 2014;23(5):554–563. doi:10.1111/JSR.12147

81. Rowland JA, Stapleton-Kotloski JR, Dobbins DL, Rogers E, Godwin DW, Taber KH. Increased Small-World Network Topology Following Deployment-Acquired Traumatic Brain Injury Associated with the Development of Post-Traumatic Stress Disorder. Brain Connect. 2018;8(4):205–211. doi:10.1089/BRAIN.2017.0556/ASSET/IMAGES/LARGE/FIGURE1.JPEG

82. Kaufeld G, Bosker HR, ten Oever S, Alday PM, Meyer AS, Martin AE. Linguistic Structure and Meaning Organize Neural Oscillations into a Content-Specific Hierarchy. Journal of Neuroscience. 2020;40(49):9467–9475. doi:10.1523/JNEUROSCI.0302-20.2020

83. Michel CM, He B. EEG source localization. Handb Clin Neurol. 2019;160:85–101. doi:10.1016/B978-0-444-64032-1.00006-0

84. Park W, Kwon GH, Kim YH, Lee JH, Kim L. EEG response varies with lesion location in patients with chronic stroke. J Neuroeng Rehabil. 2016;13(1):1–10. doi:10.1186/S12984-016-0120-2/FIGURES/4

85. Cassidy JM, Wodeyar A, Wu J, et al. Low-Frequency Oscillations Are a Biomarker of Injury and Recovery after Stroke. Stroke. Published online 2020:1442–1450. doi:10.1161/STROKEAHA.120.028932

86. Lazar RM, Antoniello D. Variability in recovery from aphasia. Curr Neurol Neurosci Rep. 2008;8(6):497–502. doi:10.1007/S11910-008-0079-X/METRICS

87. Piastra MC, Oostenveld R, Schoffelen JM, Piai V. Estimating the influence of stroke lesions on MEG source reconstruction. Neuroimage. 2022;260. doi:10.1016/J.NEUROIMAGE.2022.119422

88. Piastra MC, Van Der Cruijsen J, Piai V, et al. ASH: an Automatic pipeline to generate realistic and individualized chronic Stroke volume conduction Head models. J Neural Eng. 2021;18(4). doi:10.1088/1741-2552/ABF00B

89. Sarmukadam K, Behroozmand R. Aberrant beta-band brain connectivity predicts speech motor planning deficits in post-stroke aphasia. Cortex. 2022;155:75–89. doi:10.1016/J.CORTEX.2022.07.001

