## Supplementary material for "Functional connectivity of stimulus-evoked brain responses to natural speech in post-stroke aphasia"

Contents:

1. NBS: one-sample t-tests
2. TMIF: discrepancy across recording segments
3. Lesion overlap map
1. **NBS: one-sample t-test**

Figure S1 shows the results produced with one-sample t-tests performed with NBS on the TMIF- and EEG-networks for HC, and the results obtained from the TMIF-connectome for PWA. Topographies corresponds to group-level network components significantly above zero.

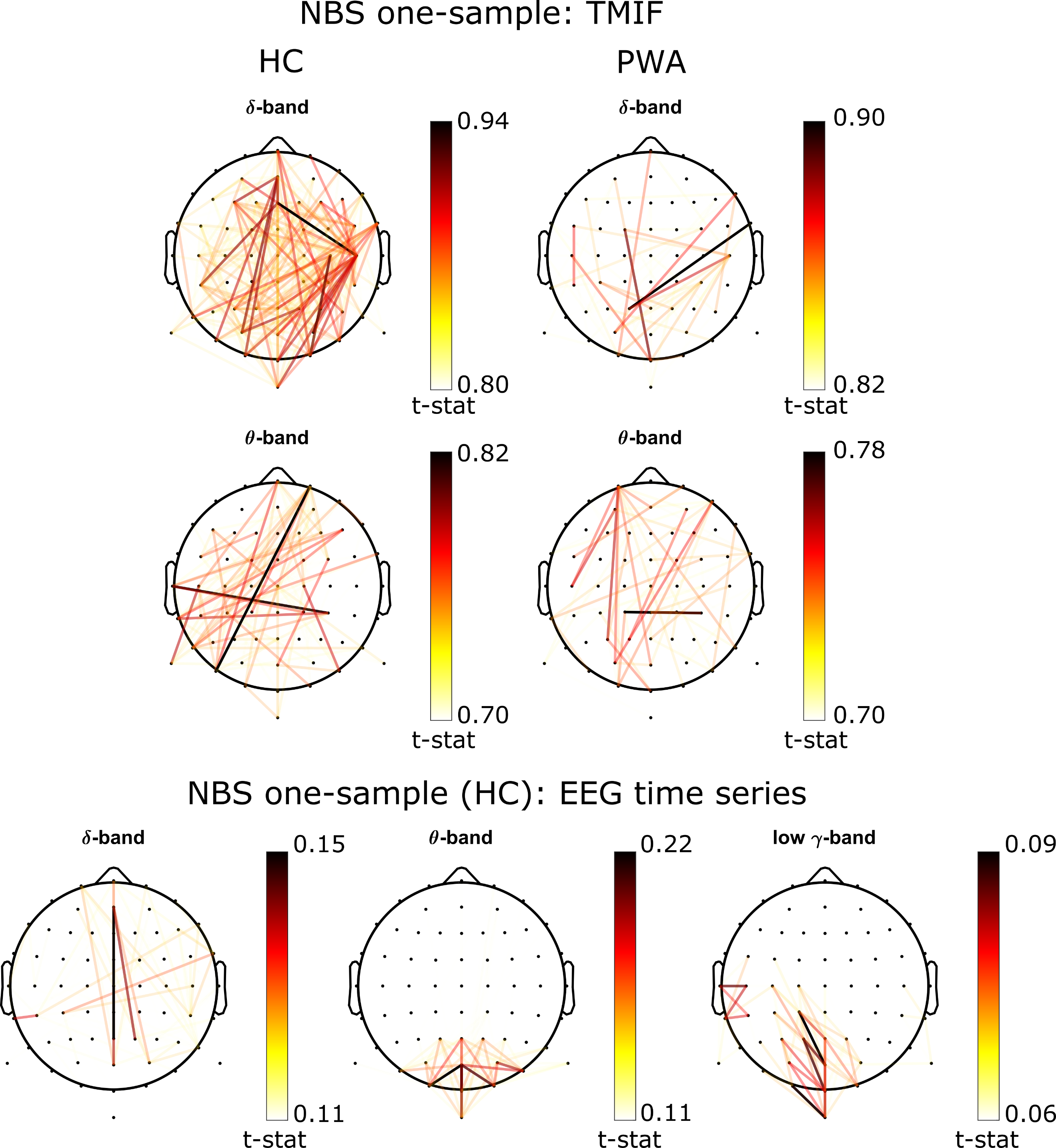

Figure S1 - NBS: one-sample t-tests. Darker colors correspond to higher t-stats (see colorbar).

2. **TMIF: discrepancy across recording segments**

For the computation of TMIFs, only the first five minutes of the experimental session was considered. As reported in the main text of this paper, this choice was aimed at excluding:

- any artifacts related to recording discontinuity across segments
- blurring effects due to visually detected discrepancies of TMIFs across recording segments (Figure S2)
- parts of the recording during which one participant fell asleep

An example of the discrepancies and shifts of TMIF waveforms across segments is shown in Figure S2, respectively for delta, theta and low-gamma bands (each frequency band from different participant).

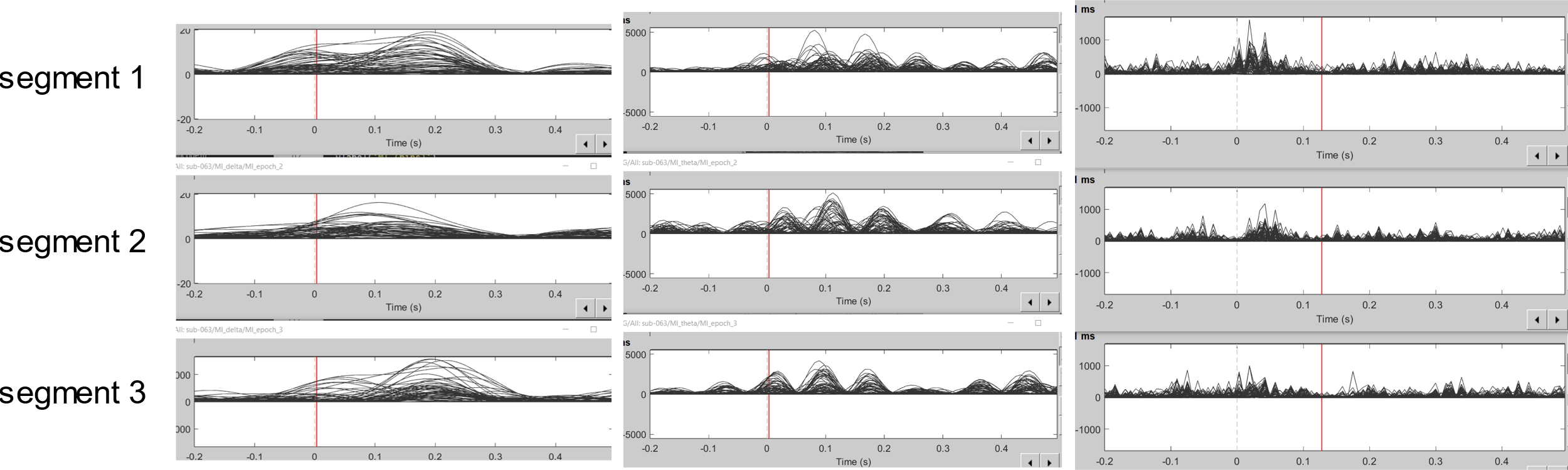

Figure S2 - Example of TMIF waveforms across three recording segments. From left to right: delta, theta, low-gamma bands (from different participants)

3. **Lesion overlap map**

To obtain an overview of the topographical distribution and the variability of the lesion across PWA, we produced a lesion overlap map using MRIcroGL toolbox (<https://www.nitrc.org/projects/mricrogl/>), shown in Figure S3. Lesioned stroke tissue was segmented on 40 T2-weigthed FLAIR (26 at the acute stage, 14 at the chronic stage) and two non-contrast enhanced computed tomography (at the acute stage). For one participant, brain scans could not be accessed, hence was excluded from the generation of the lesion overlap map. Lesions were manually delineated based on the information reported in the medical files. Despite the majority of recordings show a left-hemispheric lesion, the maximum overlap was reached for only 19 participants, reflecting a strong level of variability of lesion anatomical properties across the cohort.

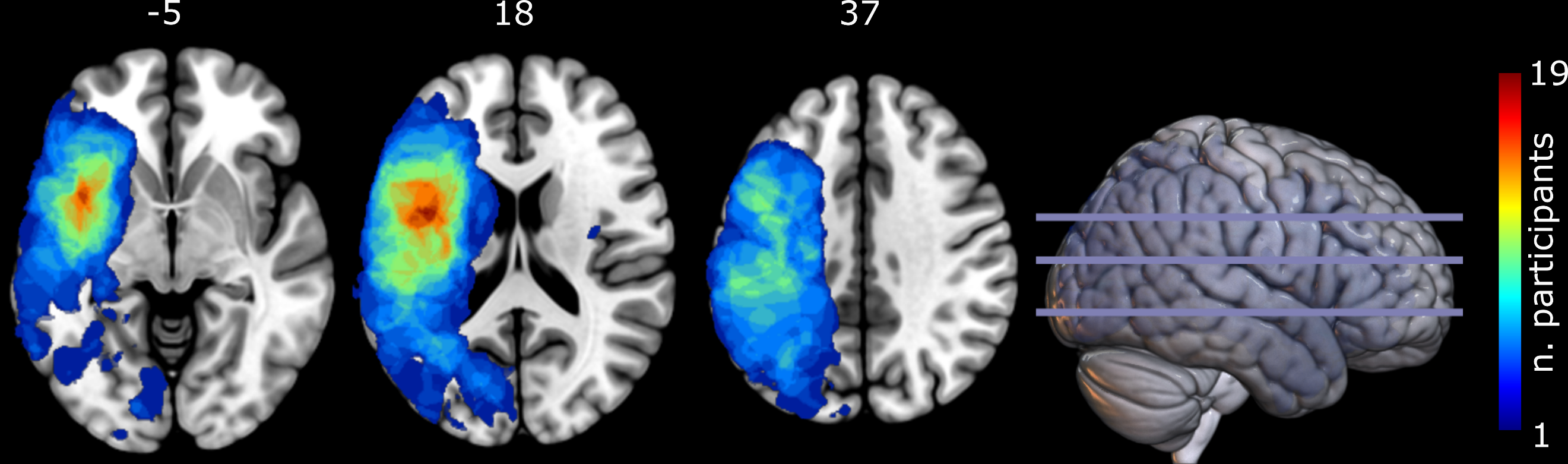

Figure S3 - Lesion overlap map. Numbers on tops correspond to the Z-layer of the MRI recording. The maximum overlap is reached for only 19 participants, over the left Rolandic area and the insula.
